# Konjac Glucomannan Improves Body Composition and Reduced Blood Cholesterol, Inflammation, and Cardiovascular Risk in Adults with Excess Weight: A Triple-Blind, Placebo-Controlled Randomized Clinical Trial

**DOI:** 10.64898/2026.04.18.26351176

**Authors:** Juan S. Escobar, Vanessa Corrales-Agudelo, O. Lucía Ortega-Sierra, N. Andrea Villota-Salazar, Diego A. Rivera, Isabel C. Pulgarín-Zapata, Mariana Hernández-Londoño, Oscar J. Lara-Guzmán, Jelver A. Sierra, Rafael Álvarez-Quintero, Juan P. Polanco, Katalina Muñoz-Durango

## Abstract

Obesity and related cardiometabolic diseases pose significant global health challenges. Konjac glucomannan, a soluble dietary fiber, has shown promise in managing these conditions. However, rigorous studies are necessary to establish its benefits on human health. We designed a parallel-arm, triple-blind, placebo-controlled RCT to test the effects of glucomannan (3 g/day, 12 weeks) on body weight and composition, lipid profile, glucose metabolism, inflammation, adipokines, intestinal permeability, gut microbiota, and fecal metabolites in 40 adults. Participants were randomly assigned to either the glucomannan or placebo group, with both groups adhering to personalized hypocaloric diets and moderate physical activity. Outcomes were analyzed as intention-to-treat using linear mixed-effect models. Irrespective of the treatment, our intervention reduced body weight (mean: -2.39 kg; 95% CI: -3.38, -1.40), BMI (−0.83 kg/m^2^; -1.15, -0.52), and waist (−2.70 cm; -3.87, -1.53). Glucomannan promoted additional benefits not obtained with the placebo, reducing body fat measured by DEXA (body fat%: -2.16%; -3.04, -1.28; VAT: -20.0 cm^2^; - 29.2, -10.8; FMI: -0.98 kg/m^2^; -1.34, -0.62), LDL (−14.1 mg/dL; -23.4, -4.9), and the atherogenic index (−0.50; -0.80, -0.21). It also diminished the Framingham score of 10-year risk of coronary heart disease (−0.370; -0.625, -0.115), C reactive protein (−1.01 mg/L; -2.18, 0.15), leptin (−2.06 ng/mL; -4.48, 0.365), and leptin/adiponectin (−0.282; -0.603, 0.040). The two treatments had similar intakes, physical activity, and adherence to the intervention. There were no adverse effects. This intervention fostered health benefits in a population at high risk of cardiometabolic diseases. Konjac glucomannan was an effective co-adjuvant for further reducing risk factors.

## INTRODUCTION

More than half a billion people around the world have obesity and other cardiometabolic affections, including cardiovascular disease and type-2 diabetes. Cardiometabolic diseases were responsible for more than 30 million global deaths in 2021.^1^ Most of these premature fatalities can be prevented by implementing strategies to reduce modifiable risk factors, such as an unhealthy diet and physical inactivity.^2^

Bioactive compounds, including antioxidants and dietary fibers from plants, have been proposed for managing cardiometabolic diseases. Studies suggest these compounds may promote weight loss, improve insulin sensitivity, and regulate blood sugar and lipids.^3^ A promising bioactive compound is obtained from konjac (*Amorphophallus konjac*), a plant of Asian origin with large starchy tubers used to obtain flour rich in glucomannan. Glucomannan is a high-molecular-weight polysaccharide (200-2000 kDa) made of β-1,4 linked glucose and mannose with a small amount of branching (∼8%) through β-1,6 glucosyl linkages.^4–6^ Glucomannan is a viscous soluble dietary fiber that can absorb up to 100 times its weight in water.^5,7^ Because human amylases and pancreases cannot completely hydrolyze β-1,4 linkages, glucomannan is non-digestible in the upper gastrointestinal tract. It passes relatively unchanged into the colon and is fermented by the gut microbiota.^8^

The European (EFSA) and American (FDA) food safety authorities recognize konjac glucomannan for its effects on weight reduction and maintaining normal blood cholesterol.^9–11^ However, recent medical reviews raised criticism about the impact of this dietary fiber,^7,12^ and two meta-analyses found conflicting results,^13,14^ likely due to the comparison of populations with different inclusion criteria, and weight losses that are of little clinical relevance.^7^ Timely, the authors of these meta-analyses called for more rigorous RCTs.^14^

Besides weight loss, questions remain about the effects of konjac glucomannan on emerging cardiometabolic risk factors. To fill these gaps in knowledge, we performed a parallel-arm, triple-blind, placebo-controlled RCT in which we evaluated the role of this fiber on obesity and well-established cardiometabolic risk factors, including blood cholesterol, blood pressure, and glycemic profile. We also assessed its effects on inflammation, adipose tissue functioning, intestinal barrier integrity, gut microbiota, and important gut microbiota-derived metabolites. Notably, this RCT employed a rigorous experimental setting with healthy diets for weight loss and moderate physical activity.

## MATERIALS & METHODS

### Products of the intervention

Naturela S.A.S. (Cumaral, Colombia) produced the two food products consumed in this study. The active product was a mix of konjac glucomannan and freeze-dried fruit, and the placebo consisted of freeze-dried fruit only. Products were individually packaged in 2.6 g sachets. Sachets of the active product provided 1 g of glucomannan. The active product and the placebo were similar in presentation and taste. Products were consumed the following way: the powder in sachets was reconstituted in 200 mL of tap water and consumed three times a day, 30 minutes before breakfast, lunch, and dinner. Products were coded directly at production with a unique code that did not refer to the product type. A pharmaceutical chemist from the research group was the only person to receive, store, and deliver the products to the dietitians, who were blind to the product type. The nutrition facts for the two products are shown in Supplementary Table S1.

### Study population and design

We recruited participants from the Obesity, Dysmetabolism, and Sports Clinical Program at Clínica Las Américas–AUNA (Medellin, Colombia), where data were collected. Physicians invited patients to participate in the study after verifying the following inclusion criteria: age 20-50 years, BMI 27.0-34.9 kg/m^2^, and at least one of the following conditions from direct examination or recent medical history: central obesity (waist circumference: >88 cm women or >102 cm men), altered blood pressure (>130/85 mm Hg), dyslipidemia (HDL cholesterol: <50 mg/dL women or <40 mg/dL men; triglycerides: >150 mg/dL), or abnormal fasting glucose (>100 mg/dL). Pregnant or lactating women, patients with medical history or recent diagnosis (less than a year) of myocardial infarction, congestive heart failure, stroke, arrhythmia, cancer, chronic renal failure, liver disorders, insomnia, duodenal ulcer, gastritis, malabsorption disorders, dysautonomia, psychiatric, autoimmune or rheumatological disorders were not included. In addition, individuals consuming metformin, steroids, anti-inflammatories, hypnotic drugs, antibiotics (last three months), antiparasitics, or laxatives were not enrolled.

We designed a parallel-arm, triple-blind, placebo-controlled RCT with a 50:50 allocation rate. The two study arms were under personalized hypocaloric diets and moderate physical activity. The study was stratified by BMI (overweight: 27.0 ≥ BMI < 30.0 kg/m^2^, obesity: 30.0 ≥ BMI < 35.0 kg/m^2^) and paired by sex (male or female) and age (± 5 years). One hundred forty-seven individuals were invited to participate in this study. Fifty-eight were assessed for eligibility. After screening, 40 eligible participants (22 women and 18 men) provided written informed consent and were gradually enrolled between February and May 2023. Participants were followed up for 12 weeks, and the follow-up period concluded in August 2023. Eligible participants were randomly assigned to two study arms and received the intended treatment: konjac glucomannan (n=20) and control (n=20). In the intervention arm, individuals consumed 3 sachets/day of the active food product (equivalent to 3 g/day of glucomannan), whereas, in the control arm, they consumed 3 sachets/day of the placebo. Each participant consumed the assigned product for 12 weeks. Participants returned empty and unconsumed sachets to the research team to verify compliance with the intervention. At baseline (week 0) and the end of the intervention (week 12), we measured primary and secondary outcomes, as described below (Figure 1).

**Figure 1.**
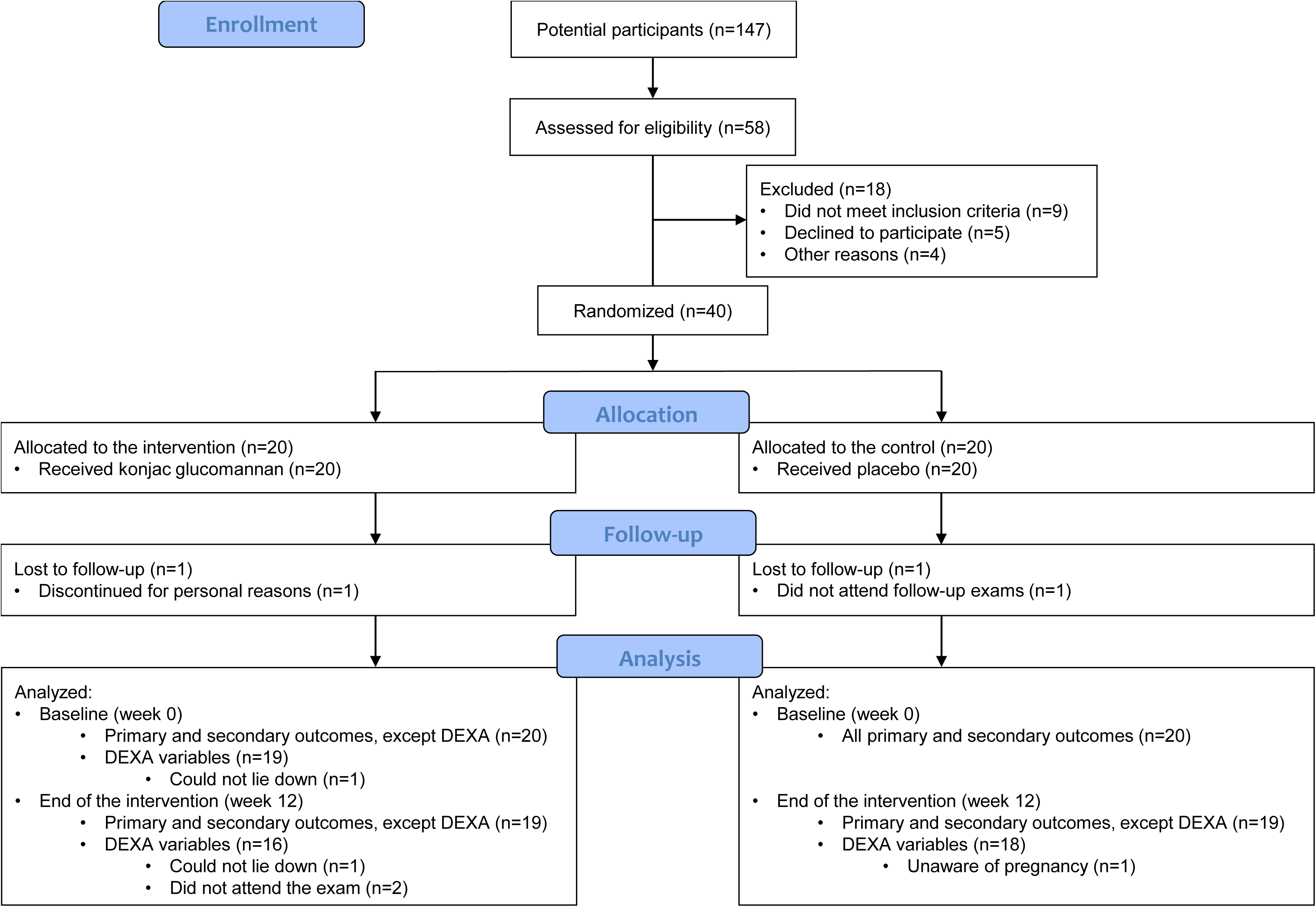
Flow diagram of the intervention.

This study was registered at the World Health Organization International Clinical Trials Registry Platform (https://trialsearch.who.int/Trial2.aspx?TrialID=RPCEC00000406).

### Randomization and blinding

Pairs of participants of similar BMI categories (overweight, obesity), age, and sex were randomly assigned to one of the two study arms. An external statistical advisor randomly allocated pairs of participants using the *sample* function without replacement in R 4.3.1.^15^ To maintain blinding, researchers, participants, and data analysts were unaware of the food product assigned to each arm.

### Sample size calculation

The sample size corresponds to a convenience sample. However, it ensures sufficient statistical power for the primary outcome (body weight). The intended sample size for this variable, assuming an expected mean difference of 2 kg, equal variance between the study arms, alpha=0.05, and power=0.8 was 4 subjects per group.

### Outcomes

At baseline (week 0) and the end of the intervention (week 12), we measured primary and secondary outcomes. As mentioned, the primary outcome was body weight. Secondary outcomes included body composition, blood pressure, biomarkers associated with metabolic status, gut microbiota (alpha diversity, beta diversity, and taxon abundance), and fecal short- and branched-chain fatty acids (SCFA and BCFA, respectively). The biomarkers associated with metabolic status were measured in blood and included lipid profile, glucose metabolism, inflammatory mediators, adipokines, and a biomarker of intestinal permeability. In addition, we assessed food intake three times during the study (weeks 3, 6, and 9) and tracked physical activity daily.

### Anthropometric measurements

We measured weight with Cardinal Detecto DR400C digital scales (Webb City, MO, USA), and height and waist circumference with Seca portable measuring rods (Hamburg, Germany). BMI was calculated as weight (kg)/height squared (m^2^). We used dual X-ray absorptiometry (DEXA) and the Hologic Discovery-W scanner system to obtain detailed body composition. We extracted the participants’ percentage of body fat, total muscular mass, fat mass index (FMI), free fat mass index (FFMI), visceral adipose tissue area (VAT), appendicular skeletal muscle mass index (ASMMI), and the android/gynoid ratio from DEXA reports.

### Blood pressure and other cardiometabolic risk factors

We obtained systolic and diastolic blood pressure with digital tensiometers. We also collected fasting venous blood for lipid profile, glucose metabolism, and other biomarkers of metabolic status. The latter included high-sensitivity C reactive protein (hs-CRP), a biomarker of systemic inflammation; the adipokines interleukin 18 (IL-18), leptin, and adiponectin, which inform about the adipose tissue functioning; the Agouti-related protein/transcript (AgRP/ART), a neuropeptide that regulates energy balance; and the lipopolysaccharide-binding protein (LBP), a biomarker of intestinal permeability. Total cholesterol, LDL, HDL, triglycerides, and fasting glycemia were measured by colorimetric enzymatic assays (cobas 701, Roche, Mannheim, Germany); fasting insulin by a chemiluminescence immunoassay (cobas E411); glycated hemoglobin (HbA1c) by high-performance liquid chromatography (Premier Hb9210); hs-CRP by a particle-enhanced immunoturbidimetric assay (cobas 502). Plasma levels of IL-18/IL-1F4, leptin, adiponectin, AgRP/ART, and LBP were measured using customized human magnetic multiplex assay kits (R&D Systems, Minneapolis, MN, USA) for Magpix (Luminex Corp., Austin, TX, USA) following the manufacturer’s recommendations. The Luminex Xponent software v4.3 was used for data analysis. We calculated the atherogenic index (total cholesterol/HDL), the homeostasis-model assessment of insulin resistance (HOMA-IR = fasting glycemia [mg/dL] ✕ fasting insulin [mUI/L]/405), and the leptin/adiponectin ratio. In addition, the Framingham risk score for the 10-year risk of coronary heart disease following risk functions derived from reference ^16^.

### Gut microbiota

Each participant collected a fecal sample, refrigerated it in portable freezers, and sent it to our laboratory within 6 hours. Samples were processed and the total DNA was extracted using Qiagen’s DNeasy Blood & Tissue kit. Total DNA was quantified using fluorometry and sent to the Microbiome Core at the University of Michigan (Ann Arbor, MI, USA) for library preparation and DNA sequencing. The V4 region of the 16S rRNA gene was amplified with primers F515 and R806.^17^ Sequencing was done on the Illumina MiSeq platform, using a MiSeq Reagent Kit v2 500 cycles. We added a mock community (ZymoBIOMICS Microbial Community DNA Standard, Zymo Research) as positive control, and ultrapure water and DNA extraction blanks as negative controls. FASTQ files were demultiplexed and processed following standard operational procedures with Mothur 1.48.0.^18^ Potentially contaminating sequences were removed with decontam 1.20.0.^19^ The table of operational taxonomic units (OTU) clustered at 97% sequence identity was analyzed using phyloseq 1.44.0.^20^ Taxonomic classification was performed using SATIVA^21^ with vetted 16S rRNA sequences^22^ from GTDB release R09RS220.^23^ The table of OTUs was rarefied to 10,000 reads/sample to homogenize the sampling effort. Alpha diversity was estimated through the Shannon diversity index, OTU richness, and Pielou’s evenness using BiodiversityR 2.15-2 (http://www.worldagroforestry.org/output/tree-diversity-analysis). Beta diversity was estimated using Aitchison distances with microViz.^24^

### Fecal SCFA and BCFA

Samples were handled and instrumental signals were acquired according to a method previously published^25^ and modified to include valeric acid (valeric acid R75054, Supelco-Belfonte, PA, USA). Aliquots of fecal samples (∼300 mg) were weighed into 10 mL magnetic screw-cap vials with PTFE septa, suitable for headspace analysis. Volatiles were sampled using a CTC Combipal 3 automated robot in HS/SPME mode, equipped with a grey fiber (Carboxen/DVB/PDMS ref. SU57329U, Supelco-Bellefonte, PA, USA). The autosampler was programmed as follows: fiber conditioning module for 5 min at 250 °C; sample equilibration for 30 min at 80 °C; analyte extraction for 25 min at 80 °C; and fiber desorption for 1 min at 250 °C at the injection port. Analytes were injected in splitless mode onto an Agilent 7890 gas chromatography (GC) system (Wilmington, DE, USA) equipped with an Agilent J&W DB-WAX capillary column (30 m, 0.25 mm, 0.250 µm). The oven temperature was programmed to start at 80 °C (5 min), increase to 100 °C for 1 min at 3 °C/min, and then increase to 250 °C for 1 min at 6 °C/min. Compound detection was performed using a 5975C mass selective detector (MS) in single ion monitoring (SIM) and scan modes. The ionization source and quadrupole temperatures were set to 230 °C and 150 °C, respectively, with an electron impact ionization energy of 70 eV. To quantify SCFA and BCFA in fecal samples, the method’s performance was evaluated for each analyte, including linearity (homoscedasticity test and residual analysis), precision (RSD < 3.0 for each analyte), and accuracy (3-way test of concentration for each analyte and 3 replicates; value = G_table_ (a=0.05; k=3; n=3) = 0.871 where G_exp_<G_table_ indicate acceptance). The detector was tuned throughout all experiments. Chromatographic peak homogeneity was verified by extracting ions from characteristic fragments to optimize resolution and peak symmetry. Data analysis was conducted using MassHunter Workstation (Agilent, Santa Clara, CA, USA). Concentrations of acetate, propionate, butyrate, isobutyrate, and valerate are expressed in µmol/g of feces.

### Diet and physical activity

At the beginning of the study (week 0), participants received personalized nutrition counseling and a personalized hypocaloric diet to lose weight healthily from expert dietitians. The designed diets considered individualized food preferences and had a minimum caloric intake (women: 1200 kcal/day, men: 1400 kcal/day). They aimed to maintain the recommended macronutrient distribution for the obese population,^26–28^ that is, 40-55% of calories from carbohydrates, 20-25% from proteins, 20-35% from fat, and 14 g of fiber per 1000 kcal/day. Food intake and adherence to diet were assessed with 24-hour dietary recalls assisted with photographs. We performed three recalls during the study, at least one on the weekend. Food items were analyzed with custom software integrating the USDA FNDDS 2017-2018 and Colombian ICBF databases to calculate energy, macronutrients, and fiber. Dietitians accompanied the participants through remote and in-person interviews to maintain motivation and adherence. We also provided participants with Xiaomi Mi Smart Band 5 to track physical activity daily. The number of steps is the most reliable variable obtained from these bands.^29^ These were monitored every day to encourage consistent physical activity.

### Statistical analysis

Statistical analyses were performed in R 4.3.1. We fit linear mixed-effects models to longitudinal outcome data with the study arm, time point, and their interaction as fixed effects, and subjects nested within recruited pairs as random effects using the *lme* function.^30^ Model assumptions of normality and homoscedasticity were assessed visually using residual plots and with Shapiro-Wilk and Fligner-Killeen tests. Variables that did not distribute normally were log-transformed or transformed using the Box-Cox power transformation.^31^ We also calculated the change between the end of the intervention and the baseline for each outcome. Least-squares means and 95% confidence intervals were calculated with emmeans.^32^ In the case of gut microbiota beta diversity, we performed permutational analyses of variance (PERMANOVA) using 10,000 permutations on Aitchison distances using the *dist_permanova* function of microViz. We also tested for differential microbial abundance using ALDEx2 1.32.0.^33^

The main statistical analyses were performed with all participants. Sensitivity analyses were performed with participants with high adherence to the intervention. Adherence was evaluated by the research team based on product consumption (returned empty sachets >80%), compliance with the diet, and physical activity.

### Ethical approval

The study was approved by the Ethical Institutional Committee for Research in Humans of CES University (Medellin, Colombia; act #166 of July 12, 2021) and Clínica Las Américas–AUNA review board (Medellin, Colombia; act #188 of July 11, 2022).

### Data and code availability

The R script and anonymized data to reproduce the results reported here are available at https://github.com/jsescobar/konjac. Raw reads from the gut microbiota characterization were deposited at the NCBI Short Read Archive under BioProject PRJNA1164676.

## RESULTS

Forty men and women were randomly assigned to two study arms and received the intended treatment in this pilot RCT: konjac glucomannan (n=20) and control (n=20). The two arms were stratified by BMI category (overweight, obesity) and paired by sex and age. Of the 40 participants who received the treatment at the beginning of the study, 38 completed it. The two participants who quit were assigned to different treatments. Detailed body composition by DEXA was performed on 39 participants at baseline and on 34 at the end of the intervention (Figure 1). The two study arms were similar in all the measured variables at baseline. As per the study design, the population consisted of young males and females in similar proportions, with excess weight, central obesity, and dyslipidemia. They were, on average, normotensive and normoglycemic (Table 1). The study ended when the last participant concluded the intervention.

**Table 1.** General characteristics of the study population at baseline. Each variable’s average value and 95% confidence intervals (lower, upper) are shown.

| Variable | Control | Konjac glucomannan |
| --- | --- | --- |
| Sex (Male: Female) | 11: 9 | 11: 9 |
| Age (years) | 36.5 (33.2, 39.9) | 37.2 (33.9, 40.6) |
| BMI (kg/m<sup>2</sup>) | 30.7 (29.7, 31.8) | 30.9 (29.8, 31.9) |
| Waist circumference (cm) | 94.6 (90.4, 98.8) | 93.7 (89.5, 97.9) |
| Systolic blood pressure (mm Hg) | 122 (116, 127) | 125 (120, 130) |
| Diastolic blood pressure (mm Hg) | 78.8 (74.9, 82.7) | 83.5 (79.6, 87.4) |
| HDL cholesterol (mg/dL) | 46.7 (41.3, 52.0) | 44.9 (39.5, 50.3) |
| Triglycerides (mg/dL) | 144 (113.6, 175) | 146 (115.5, 176) |
| Fasting glycemia (mg/dL) | 89.7 (86.2, 93.1) | 85.8 (82.4, 89.3) |

### Body weight and composition

We observed significant improvements in anthropometry, including reductions in body weight (primary outcome), BMI, and waist circumference in the control and konjac glucomannan groups after 12 weeks of intervention (Table 2). All participants, irrespective of the treatment, reduced body weight by about 3% at the end of the intervention compared to the baseline (mean: -2.39 kg, 95% CI: -3.38, -1.40). This represented a mean BMI change of -0.83 kg/m^2^ (95% CI: -1.15, -0.52) and a mean waist circumference change of -2.70 cm (95% CI: -3.87, -1.53). Body composition analyzed by DEXA indicated that participants consuming konjac glucomannan experienced significantly greater reductions in VAT, body fat percentage, and FMI than participants in the control arm. Participants in the control experienced statistically significant within-arm losses in lean mass at the end of the intervention compared to the baseline. This included total muscular mass, FFMI, and ASMMI. These changes were not statistically significant in the between-treatment comparisons. On the other hand, participants consuming konjac glucomannan significantly reduced the android/gynoid ratio in the within-arm comparison between the end of the intervention and the baseline. Importantly, none of the changes above were explained by differences in intake or physical activity between the two study arms (Supplementary Table S2).

**TABLE 2.**
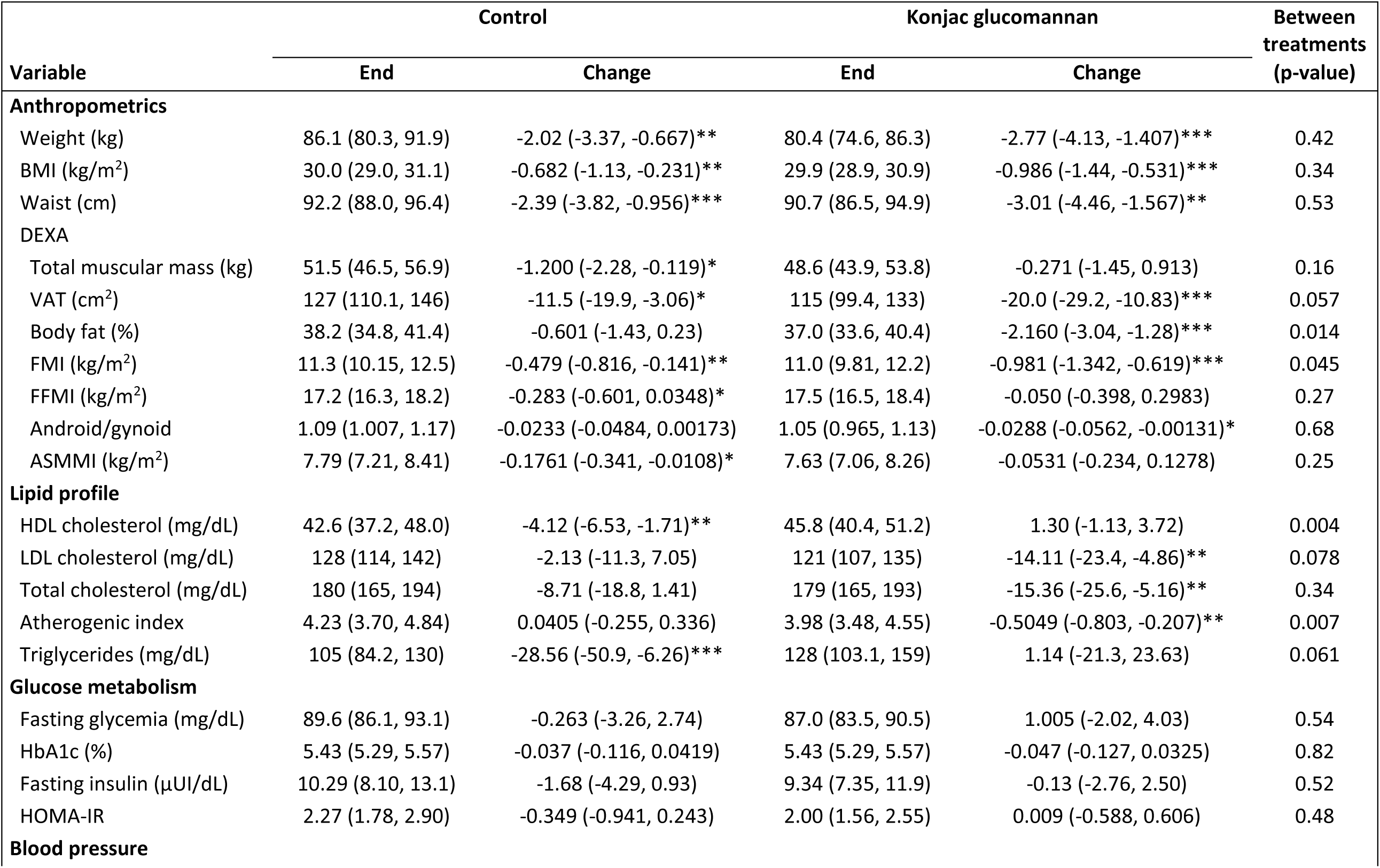
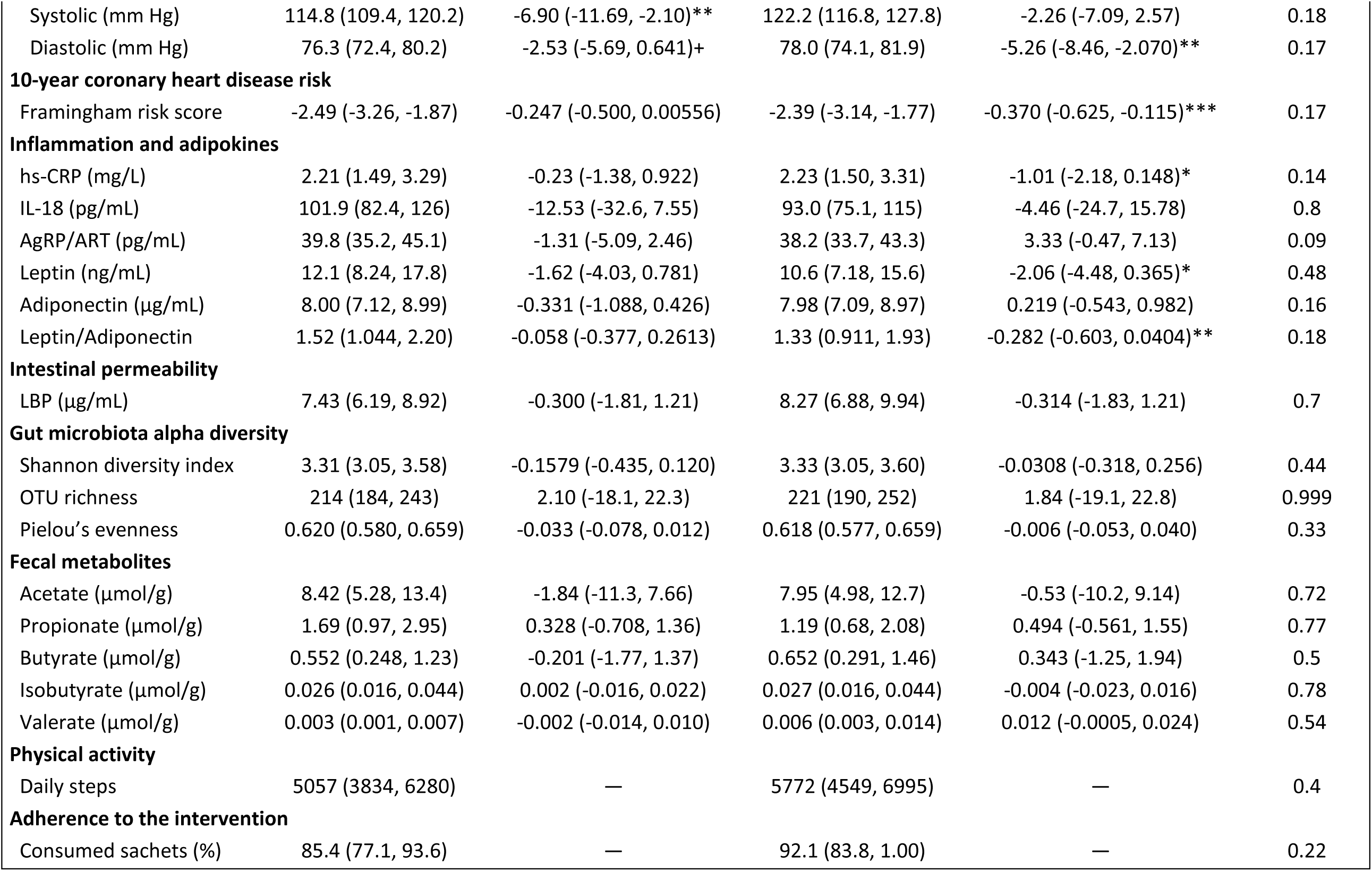
Effect of the intervention in primary and secondary outcomes. Average and 95% confidence intervals (lower, upper) are shown at the end of the intervention (week 12) and for the change between the baseline and the end of the intervention. Within-treatment effects are denoted as + = p<0.10; * = p<0.05; ** = p<0.01; *** = p<0.001.

### Lipid profile and glucose metabolism

Participants in the two study arms significantly differed in their blood cholesterol changes between the end of the intervention and the baseline. This included differences in HDL and the atherogenic index. Within-arm analysis indicated that participants who consumed konjac glucomannan significantly reduced LDL, total cholesterol, and atherogenic index, whereas participants in the control arm significantly reduced HDL and triglycerides. None of the two treatments promoted consistent changes in the glycemic profile (Table 2).

### Blood pressure and 10-year risk of coronary heart disease

We observed no differences between the two study arms in the changes observed in blood pressure or the Framingham score between the end of the intervention and the baseline. According to the Framingham risk score, of the 38 participants who completed the trial, 29 diminished their 10-year risk of coronary heart disease and nine increased it. All participants maintained a risk below 10% throughout the study. Within-arm analyses indicated that participants in the control arm significantly reduced their systolic blood pressure. Participants consuming konjac glucomannan significantly diminished their diastolic blood pressure and the 10-year risk of coronary heart disease (Table 2).

### Inflammation and adipokines

To delve into the physiology of the konjac glucomannan-related changes in anthropometry and blood cholesterol, we measured additional variables related to inflammation, the functioning of the adipose tissue, and appetite regulation. Within-arm analyses showed that participants consuming konjac glucomannan significantly reduced hs-CRP, leptin, and the leptin/adiponectin ratio. These changes were not observed in placebo participants (Table 2).

### Intestinal barrier, gut microbiota, and gut microbiota-derived metabolites

We also explored whether konjac glucomannan promoted changes in the gastrointestinal tract, by shifting the permeability of the intestinal barrier, measured through the plasmatic levels of LBP, gut microbiota, and the fecal excretion of SCFA and BCFA. None of the changes between the baseline and the end of the intervention in these outcomes differed between the two study arms (Table 2). PERMANOVA on Aitchison distances indicated that microbial composition was mostly explained by the participant from whom the sample was taken (R^2^=0.78, p<0.0001), whereas the treatment effect was marginally significant and described a smaller proportion of the variance (R^2^=0.02, p=0.05). Indeed, Aitchison distances between baseline and end-of-the-intervention samples from the same subject were significantly smaller than between samples from different subjects (Figure 2). In terms of microbial abundance, no OTU, phylum, class, order, family, genus, or species was consistently and significantly over- or under-detected between the baseline and the end of the intervention after adjusting p-values for multiple comparisons (Supplementary Table S3).

**Figure 2.**
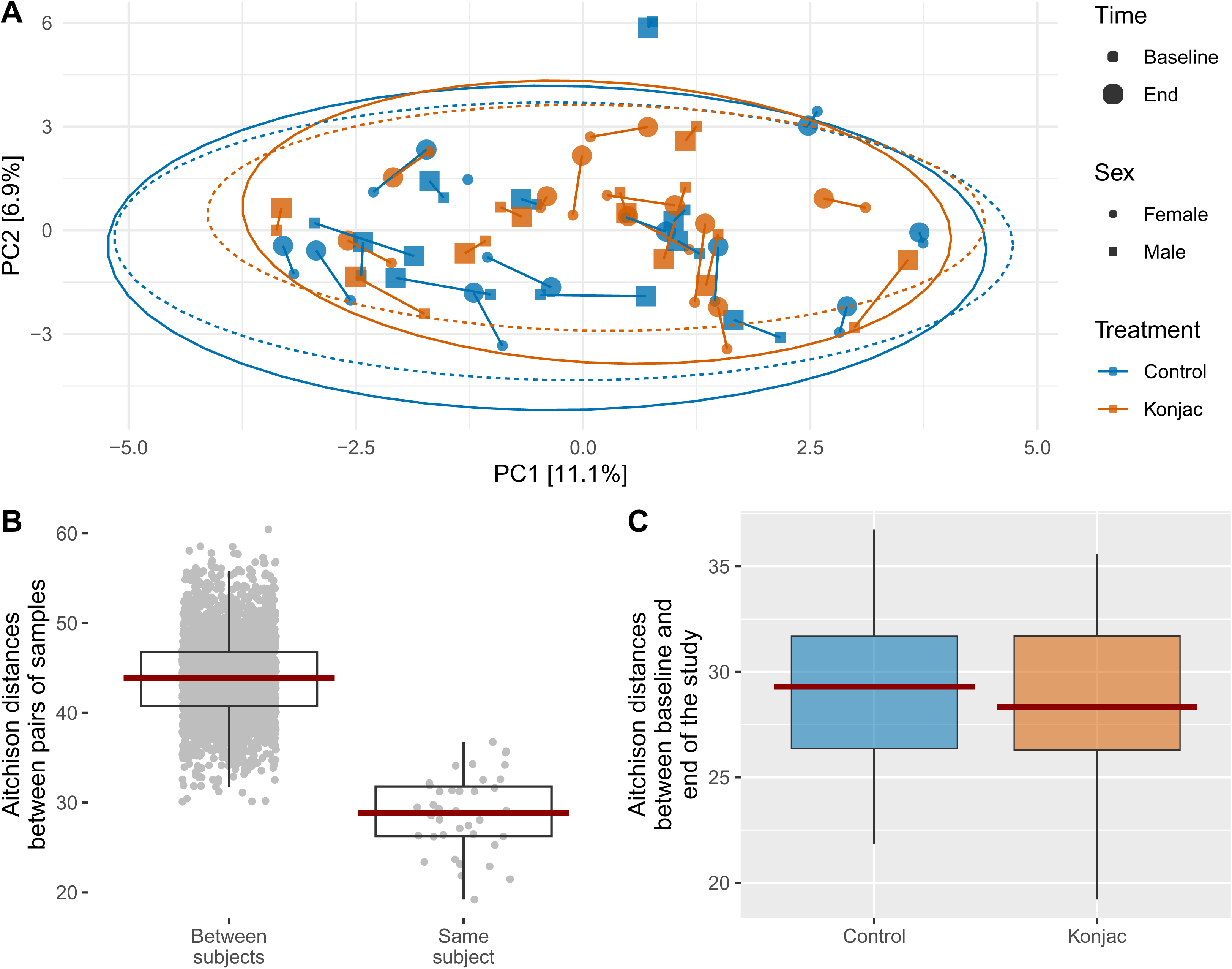
Effect of the intervention on the gut microbiota. (A) Principal component analysis based on gut microbiota beta diversity (Aitchison distances). Each point represents the gut microbiota composition at baseline (small dots) or the end of the intervention (big dots). The line uniting two points indicates the change following the intervention. The ellipses calculated for each treatment and time point encompass 95% of the variance. Control=blue; intervention=orange. (B) Aitchison distances comparing pairs of samples of the same subject or from different subjects. (C) Aitchison distances between the baseline (week 0) and the end of the intervention (week 12) for each treatment. The lower and upper hinges of the box-and-whisker plots correspond to the first and third quartiles, the arithmetic mean is shown as a horizontal red bar.

### Sensitivity analyses

We repeated analyses after excluding participants with low adherence to the intervention. Results were similar in this smaller set of highly adherent participants than in the whole cohort (Supplementary Table S4).

## DISCUSSION

In this parallel-arm, triple-blind, placebo-controlled RCT, we delved into the effects of konjac glucomannan on human health by analyzing variables related to obesity and other cardiometabolic diseases under stringent experimental conditions, including personalized healthy diets for weight loss and moderate physical activity. We found that adherence to this intervention for 12 weeks, irrespective of the food product consumed, reduced body weight, BMI, and waist. Supplementation with konjac glucomannan promoted additional benefits not obtained with the placebo, reducing body fat and blood cholesterol, and shifting levels of adipose tissue mediators. The intervention did not promote changes at the gastrointestinal level (*i.e.*, intestinal barrier permeability, gut microbiota, or fecal metabolites), and did not have adverse effects.

Different studies have tested the impact of konjac glucomannan on body weight and body fat in overweight and obese subjects. While we found significant reductions in body weight, BMI, and waist circumference in both study arms, meaning that the beneficial effects of our intervention on these variables were due to improved diet and physical activity, we observed that participants consuming konjac glucomannan reduced the body fat percentage, FMI, and VAT significantly more than participants in the control arm. Various double-blind, placebo-controlled RCTs with sample sizes, intervention durations, and doses similar to ours found that subjects consuming konjac glucomannan had body weight losses significantly higher than those consuming a placebo. Indeed, EFSA acknowledged a cause-and-effect relationship between the consumption of konjac glucomannan and body weight reduction in the context of an energy-restricted diet based on several of these studies.^9^ Walsh et al.^34^ found that obese subjects consuming 3 g/day of glucomannan for 8 weeks had significantly higher weight loss than those consuming a placebo. Similar results were found by Vita et al.^35^ with 50 subjects with obesity followed for 3 months and consuming 4 g/day of konjac glucomannan, and by Cairella and Marchini^36^ with 30 overweight women consuming the same dose for 60 days. Birketvedt et al.^37^ studied 52 healthy overweight individuals consuming 1.24 g/day of glucomannan for 5 weeks and found they had significantly higher weight loss than those consuming a placebo. In a single-blind, placebo-controlled, crossover trial with 20 men and women with type-2 diabetes, Chearskul et al.^38^ reported that 3 g/day of konjac glucomannan for 4 weeks produced significant BMI reductions. In a larger parallel-arm, double-blind, placebo-controlled RCT with 200 patients under energy-restricted diets, Salas-Salvadó et al.^39^ found that a mixed fiber dose including 3 g of *Plantago ovata* husk and 1 g of konjac glucomannan for 16 weeks tended to produce higher weight loss than the placebo. Kaats et al.^40^ analyzed 83 overweight adults for 60 days finding that those compliant or partially compliant with the intervention (3 g/day of konjac glucomannan) reduced body weight, percentage of body fat, and fat mass more than participants under the placebo. More recently, in a double-blind, placebo-controlled, crossover study 32 individuals with metabolic syndrome who received a daily serving of 400 g of noodles made with 4 g of konjac glucomannan experienced significant decreases in body weight, BMI, and waist circumference after 4 weeks of intervention.^41^ Fernandes et al.^42^ performed a double-blind, placebo-controlled RCT in which they followed 42 young adults consuming two candies per day containing 250 mg of konjac glucomannan, and found a reduction in waist circumference.

While these RCTs were performed in short periods and considered a small number of participants, evidence supports the beneficial role of konjac glucomannan on obesity. The mechanism that explains this effect involves the fiber’s rheological properties. Glucomannan is a soluble fiber that hydrates in the stomach, forming a hydrocolloid that delays gastric emptying, reduces bowel transit, and induces satiety, subsequently decreasing energy intake.^8,43–45^ Fecal energy loss could also explain reductions in body weight because soluble fibers reduce fat and protein absorption.^7^

In addition to the benefits in anthropometry, konjac glucomannan significantly improved blood cholesterol, especially HDL, LDL, and atherogenic index, while producing no effect on the glycemic profile, likely because our participants were mostly normoglycemic. In 2009, EFSA acknowledged a cause-and-effect relationship between the consumption of konjac glucomannan and the reduction of blood cholesterol.^46^ More recently, a systematic review and meta-analysis of 12 studies and 370 subjects showed that konjac glucomannan was responsible for significant reductions in LDL (−10%) and non-HDL cholesterol (−7%).^47^ These values align with the average reductions observed in our study: -9.2% (95% CI: -15.2, -3.1) for LDL and -7.2% (95% CI: -12.0, -2.5) for total cholesterol. The viscosity of konjac glucomannan would be responsible for these effects, as it diminishes fat absorption,^48^ suppresses the synthesis of hepatic cholesterol and increases the synthesis of bile acids, their reabsorption, and fecal excretion.^49,50^

Collectively, our results on body composition and blood cholesterol indicate that supplementation with 3 g/day of konjac glucomannan for 12 weeks, in the context of an energy-restricted diet and moderate physical activity (∼5000 steps/day) was effective in improving well-established cardiometabolic risk factors and reducing the risk of coronary heart disease, as per the analysis of the Framingham risk score.

Our intervention also promoted shifts in emerging biomarkers of cardiometabolic disease. Within-treatment analyses indicated that participants consuming konjac glucomannan reduced levels of hs-CRP, leptin, and the leptin/adiponectin ratio. High-sensitivity CRP is a marker of inflammation associated with an increased risk of cardiovascular disease and diabetes.^51^ Observational studies, RCTs, and meta-analyses have consistently shown that increased dietary fiber intake significantly reduces CRP levels.^52–56^ The mechanisms by which dietary fiber reduces inflammation may involve weight loss, changes in insulin and glucose metabolism, and alterations in the secretion of adipokines.^52,56^ Leptin and adiponectin are two adipokines that regulate feeding, energy balance, insulin sensitivity, and inflammation. The leptin/adiponectin ratio has emerged as a biomarker in assessing the risk of cardiovascular events. This ratio is considered more indicative of cardiovascular risk than leptin or adiponectin levels alone^57^ due to its association with metabolic and inflammatory processes that contribute to atherosclerosis and other cardiovascular conditions. The leptin/adiponectin ratio is considered an informative indicator of adipose tissue dysfunction in patients with metabolic syndrome and obesity.^58,59^ An increased ratio correlates with higher cardiovascular risk due to its association with insulin resistance and lipid disorders.^60^

Dietary fibers resist digestion in the upper gastrointestinal tract and are fermented in the colon by gut microbes, increasing the production of SCFA and BCFA. In agreement with previous studies, our intervention produced personalized compositional changes in the gut microbial community, with no consistent effect contributed by the treatment. High inter-individual gut microbiota variation may be due to human genetics, habitual diet, health conditions, transit time, and stool consistency.^61^ The fact that our participants’ diets were based on individualized food preferences contributed to idiosyncratic gut microbiota responses. Inter-individual variability in gut microbiota composition has been shown to overwhelm variation due to nutritional interventions.^62^ Our intervention did not produce changes in the fecal excretion of SCFA or BCFA. This could be because there were no differences in macronutrient or fiber intake between the two treatments.

The results of this study give insights into the reduction of cardiometabolic disease risk using natural products. However, it has several limitations. First, it considered a small number of subjects. It was designed with a sample size that guaranteed sufficient statistical power for the primary outcome, but not for secondary outcomes. As such, it should be viewed as a pilot study that puts forward hypotheses about the potential benefits of konjac glucomannan on human health. Also, its duration was limited to 12 weeks. A longer follow-up would be necessary to verify that the observed effects are stable in time. Finally, some clinical outcomes were of small magnitude in the study period. Konjac glucomannan should be viewed as a co-adjuvant for reducing cardiometabolic risk in the context of a healthy lifestyle.

## CONCLUSION

Our controlled intervention with healthy diets for weight loss and moderate physical activity reduced several cardiometabolic disease risk factors. Konjac glucomannan was an effective co-adjuvant for further promoting health benefits, including shifts in body fat, blood cholesterol, inflammation, and adipokines.

## ACKNOWLEDGMENTS

We are grateful to the study participants for their commitment to the intervention. We thank the staff at Clínica Las Américas–AUNA for the logistics and help. We also thank Ricardo Rosero for his insights into the project conception, and Rubén Manrique for his contribution to the study design and randomization of participants. We are indebted to Andrea Bolívar at Naturela S.A.S. for product manufacturing.

## AUTHOR CONTRIBUTION

**J.S.E.:** conceptualization, data curation, formal analysis, writing-original draft; **V.C.-A.:** conceptualization, data curation, formal analysis, writing-review & editing; **O.L.O.-S.:** conceptualization, data curation, formal analysis, writing-review & editing; **N.A.V.-S.:** conceptualization; data curation, formal analysis, methodology, writing-review & editing; **D.A.R.:** formal analysis, methodology; **I.C.P.-Z.:** formal analysis, writing-review & editing; **M.H.-L.:** data curation, formal analysis; **O.J.L.-G.:** conceptualization, data curation, formal analysis, methodology, writing-review & editing; **J.A.S.:** conceptualization, data curation, formal analysis, methodology, writing-review & editing; **R.A.-Q.:** data curation, formal analysis, writing-review & editing; **J.P.P.:** conceptualization, supervision, writing-review & editing; **K.M.D.:** conceptualization, funding acquisition, project administration, supervision, writing-review & editing.

## ABBREVIATIONS

ASMMI: appendicular skeletal muscle mass index
AgRT/ART: Agouti-related protein/transcript
DEXA: dual X-ray absorptiometry
FFMI: free fat mass index
FMI: fat mass index
HOMA-IR: homeostasis model assessment-insulin resistance
HbA1c: glycated hemoglobin
IL-18: interleukin 18
LBP: lipopolysaccharide-binding protein
OTU: operational taxonomic unit
RCT: randomized clinical trial
VAT: visceral adipose tissue area
hs-CRP: high-sensitivity C reactive protein.

